# Exploring dynamic alpha band connectivity in Parkinson’s disease: A novel approach to postural control assessment using the BioVRSea paradigm

**DOI:** 10.1101/2025.04.06.25325331

**Authors:** Federica Pescaglia, Lorena Guerrini, Carmine Gelormini, Romain Aubonnet, Gylfi Örn Thormar, Giorgio Di Lorenzo, Halldór Jónsson, Mahmoud Hassan, Paolo Gargiulo

## Abstract

**Objective:** Parkinson’s Disease (PD) is a neurological disorder characterized by impaired postural control (PC) and balance issues. To date, few studies have explored the relationship between brain activity and responses during specific tasks designed to challenge balance in individuals with PD. Our exploratory research employs an innovative paradigm to assess PC by integrating virtual reality (VR) and electroencephalography (EEG).

**Approach:** In the study, 20 individuals diagnosed with PD who self-reported postural instability participated in the BioVRSea paradigm. This paradigm tested their PC using visuomotor stimuli and collected EEG signals to assess brain responses throughout the experiment. The results of the Parkinson’s group were compared with those of 22 age-matched healthy controls (CTR). From functional connectivity between brain regions, we employed novel techniques that use clustering algorithms to identify brain network states (BNSs). These BNSs define brain dynamics and can be compared with resting-state networks (RSNs) to further explore and identify neural alterations in individuals with PD.

**Main Results:** Six distinct BNSs were identified, with the dorsal attention network (DAN) dominant in five states. A significant reduction in the occurrence of BNS2 (p=0.005) was observed in PD patients during the PRE movement and visuomotor (MOV) phases compared to CTR. This reduced occurrence of BNS2 suggests impaired visuomotor integration in PD patients during PC tasks. DAN dominance highlights its crucial role in maintaining attentional control during the task.

**Significance:** The findings of this study highlight the potential of using brain dynamics as a biomarker of neural dysfunction in PD, especially during specific PC tasks. Altered BNSs, particularly in networks associated with attention and sensorimotor integration, reveal key neural deficits related to PD.

## Introduction

### Parkinson’s Disorder

Parkinson’s disease (PD) is a widespread neurodegenerative condition characterized by tremors, stiffness, slowed movements, and balance problems [1]. In addition to these motor symptoms, individuals with PD often experience non-motor symptoms such as cognitive decline, mood disorders, difficulty with speech, and swallowing. These symptoms often progress slowly, affecting the ability to perform daily activities and significantly affecting the quality of life. As the brain struggles to respond accurately to external stimuli in PD, motor tasks become more challenging, leading to increased effects of neurodegeneration [2, 3]. The diagnosis of PD is a significant clinical challenge due to the overlap of motor and non-motor symptoms with other neurological deficits [4]. Early signs can often be mistaken for typical age-related changes, complicating the diagnostic process. Since there are no definitive biomarkers for PD, the diagnosis is primarily based on clinical evaluations along with a range of diagnostic tools. These include gait analysis to assess motor function, brain imaging techniques like MRI or PET scans to detect structural or functional changes in the brain, and electroencephalography (EEG) to monitor electrical activity. These approaches are essential in helping clinicians distinguish PD from other conditions with similar symptoms, although an accurate diagnosis can still be challenging. Studies on quantitative gait analysis have shown that measures like stride length, gait speed, and variability in step timing can effectively distinguish PD from other movement disorders [5]. This technique is especially valuable for monitoring the progression of PD and assessing the effectiveness of treatments, such as levodopa [6]. On the other hand, brain imaging techniques offer critical insight into structural and functional anomalies associated with PD. Magnetic resonance imaging (MRI) is used primarily to exclude other neurological conditions and can reveal subtle changes in brain regions, such as the substantia nigra [7]. However, it is limited in its ability to detect early neurodegenerative changes associated with PD. Functional imaging techniques, such as Positron Emission Tomography (PET) and Single Photon Emission Computed Tomography (SPECT), provide more specific information by visualizing the dopaminergic deficits common in PD [8, 9]. Emerging techniques, such as near-infrared spectroscopy (NIR), are also gaining attention due to their potential to assess cortical hemodynamics and oxygenation patterns in PD. NIR offers a non-invasive means of examining brain function, with recent studies suggesting it could be a useful adjunct to other imaging modalities in diagnosing PD [10].

### EEG as a diagnostic tool

EEG has been emerged as a complementary diagnostic tool for PD, showing great potential due to its unique advantages in terms of temporal resolution [11–13]. Compared to more expensive imaging methods like PET or MRI, EEG is not only cost-effective but also portable, making it suitable for use in various clinical environments. Additionally, it is non-invasive, requiring no exposure to radiation or contrast agents. EEG records brain activity with millisecond precision, enabling the monitoring of fast neural dynamics linked to motor control and cognition. This real-time insight is particularly valuable in PD, where the progressive degeneration of dopaminergic neurons disrupts brain-body communication. EEG can detect these disruptions as they occur, potentially revealing alterations in neural activity patterns before symptoms like tremors or bradykinesia emerge. This capability to capture early neural changes highlights the promise of EEG in identifying early-stage or preclinical markers of PD [14–16]. Moreover, its ability to examine changes in brain connectivity and resting-state networks, often disrupted in PD, provides an added layer of diagnostic capability.

### EEG in postural tasks

Furthermore, EEG is increasingly being recognized for its role in assessing PC in neurological disorders [17–19]. PC refers to the complex integration of sensory inputs, motor responses, and neural processes that help maintain balance and stability [20, 21]. In PD, the progressive degeneration of dopaminergic neurons disrupts these mechanisms, leading to postural instability, which significantly increases the risk of falls and injuries [22, 23]. Studies have shown that PD patients demonstrate irregular brainwave patterns during PC tasks. Recent research has linked alpha band activity in EEG to various aspects of PC, including balance maintenance and postural adjustments [24–26]. For instance, increases in alpha power are associated with improved postural stability [27, 28], while decreases indicate greater attention demands or instability [29,30]. Alpha coherence between brain areas has been shown to play a role in sensory processing and motor planning, both of which are important for maintaining balance [31, 32]. The integration of sensory information becomes particularly challenging in PD, where patients exhibit a reduced capacity to combine input from the body and environment. EEG analyses reveal disrupted connectivity and decreased synchronization among brain regions involved in sensory integration and motor planning, such as the sensorimotor cortex and parietal areas [33]. These dysfunctions significantly contribute to the postural instability characteristic of PD.

### BioVRSea

Most EEG investigations in PD focus on resting-state and static tasks, leaving a gap in understanding brain activity during dynamic tasks requiring continuous postural adjustments. Our study introduces a new paradigm called BioVRSea, a system designed to stimulate PC responses using a virtual reality (VR) environment and moving platform. Previous studies have demonstrated the effectiveness of BioVRSea as a tool for evaluating PC [34, 35]. Moreover, this setup has proven to be a promising tool for developing predictive models for various conditions [36, 37], including individuals who have experienced concussions or suffer from motion sickness [38]. A recent study using BioVRSea in PD [39] has revealed significant differences in theta and alpha power spectra bands between early-stage PD patients and healthy controls, highlighting its potential to offer deeper insights into the neural mechanisms behind postural instability in PD.

### Brain Network States (BNSs) in PD

The evaluation of PC response using a dynamic approach on EEG, such as the identification of brain network states (BNSs), has recently demonstrated significant potential [40]. In recent years, advanced EEG techniques have been developed to analyze functional connectivity by reconstructing brain sources from scalp signals and identifying brain networks. The BNSs refer to specific patterns of connectivity in the brain that change to adapt to various demands and environments [41, 42]. In PC, maintaining balance and stability requires the integration of sensory information, motor planning, and cognitive processes, all of which are influenced by the activity of distinct brain networks. Altered BNSs in PD have been identified through fMRI and EEG, showing disrupted connectivity in critical networks related to motor control, attention, and sensorimotor integration [43]. These alterations affect regions like the supplementary motor area and primary motor cortex during both rest and motor tasks. Additionally, the altered BNS configurations in PD impact cortical-striatal circuits involved in motor planning. These results reflects how the neurodegenerative disease affects the dynamic reconfigurations of the brain and presents potential biomarkers for diagnosis and tracking progression. Established networks, including the default mode network, the auditory network, the salience network, the executive control network, and the sensorimotor cortex, have been linked to specific cognitive tasks and postural adjustments [44, 45]. These networks are crucial for balance-related processes, highlighting their significance in maintaining stability and coordinating motor function [46, 47]. Furthermore, disruptions in BNS can lead to impaired PC, which is particularly evident in individuals with neurodegenerative conditions such as PD. By studying BNSs in relation to PC, research can identify potential biomarkers for diagnosing balance disorders and tracking disease progression.

### Innovations in our study

The study introduces several innovations for the evaluation of early-stage PD, in particular during a complex PC task. A key advancement is the use of dynamic EEG analysis to identify brain networks in PD. These BNSs provide a deeper understanding of the functional organization of the brain during our experiment, offering insights into how the brain adapts to different tasks and conditions. While there is no universally defined optimal method for processing EEG data to identify brain networks, we employed the most advanced methodologies, which have shown the best performance in previous studies [48]. This approach allows for a more precise analysis of brain connectivity, enhancing our understanding of PD progression and treatment responses. The nature of this work is exploratory, as we focus on investigating a specific frequency range, the alpha band. No previous study has focused on PD during a specific visuomotor task while investigating brain dynamics. By analyzing BNSs during the BioVRSea paradigm, a task that is both cognitively and motorically demanding, we aim to identify neurophysiological markers of PD. Our findings could contribute to early-stage diagnosis by detecting changes in brain connectivity and motor control mechanisms associated with PD.

## Materials and methods

### Participants

The study involved 42 participants: 20 PD patients (54–82, mean age: 65.35 years, Male =60%) and 22 age-matched healthy controls (CTR) individuals (55–72, mean age: 62.78 years, Male =68.18%). As shown in Tab.1, neither the t-test for age nor the chi-squared test for gender revealed significant differences, suggesting a uniformity in the population characteristics across both groups. Participants in the PD group had a diagnosis of early-stage PD, and they were under levodopa medication. The Hoehn and Yahr (H/Y) scale [49] was used to classify Parkinson’s patients, with stages ranging from 1 to 3. After a medical review, 11 patients showed no signs of cognitive decline, while the remaining patients were classified as having mild cognitive impairment (MCI). None were diagnosed with Parkinson’s dementia, and none exhibited obvious motor symptoms. Every participant successfully completed the full experiment and responded to both a pre-experiment and a post-experiment questionnaire. From these responses, we did not register any cognitive issues or impairments.

**Table 1.** Age and gender distribution in PD and CTR groups. No significant differences were found (t-test for age, chi-squared test for gender).

|  | PD (n=20) | CTR (n=22) | P value |
| --- | --- | --- | --- |
| **Age** |  |  | 0.182 |
| Mean ± SD | 65.4 ± 4.50 | 62.8 ± 4.27 |  |
| Min-Max | 54-82 | 55-72 |  |
| **Gender (%)** |  |  | 0.817 |
| Female | 8 (40%) | 12 (54.5%) |  |
| Male | 12 (60%) | 10 (45.5%) |  |

### BioVRSea paradigm

The multi-biometric BioVRSea setup simulates a sea environment where the subject is positioned on a small virtual boat synchronized with a moving platform. The innovative system, shown in Fig.1, integrates EEG with VR to induce and assess PC through various visuomotor stimulations. This environment is designed to create a sensation of imbalance, prompting the participant to engage in postural strategies to maintain equilibrium. The movement of the platform introduces a dynamic balance challenge, which becomes particularly pronounced towards the end of the experiment.

**Fig 1.**
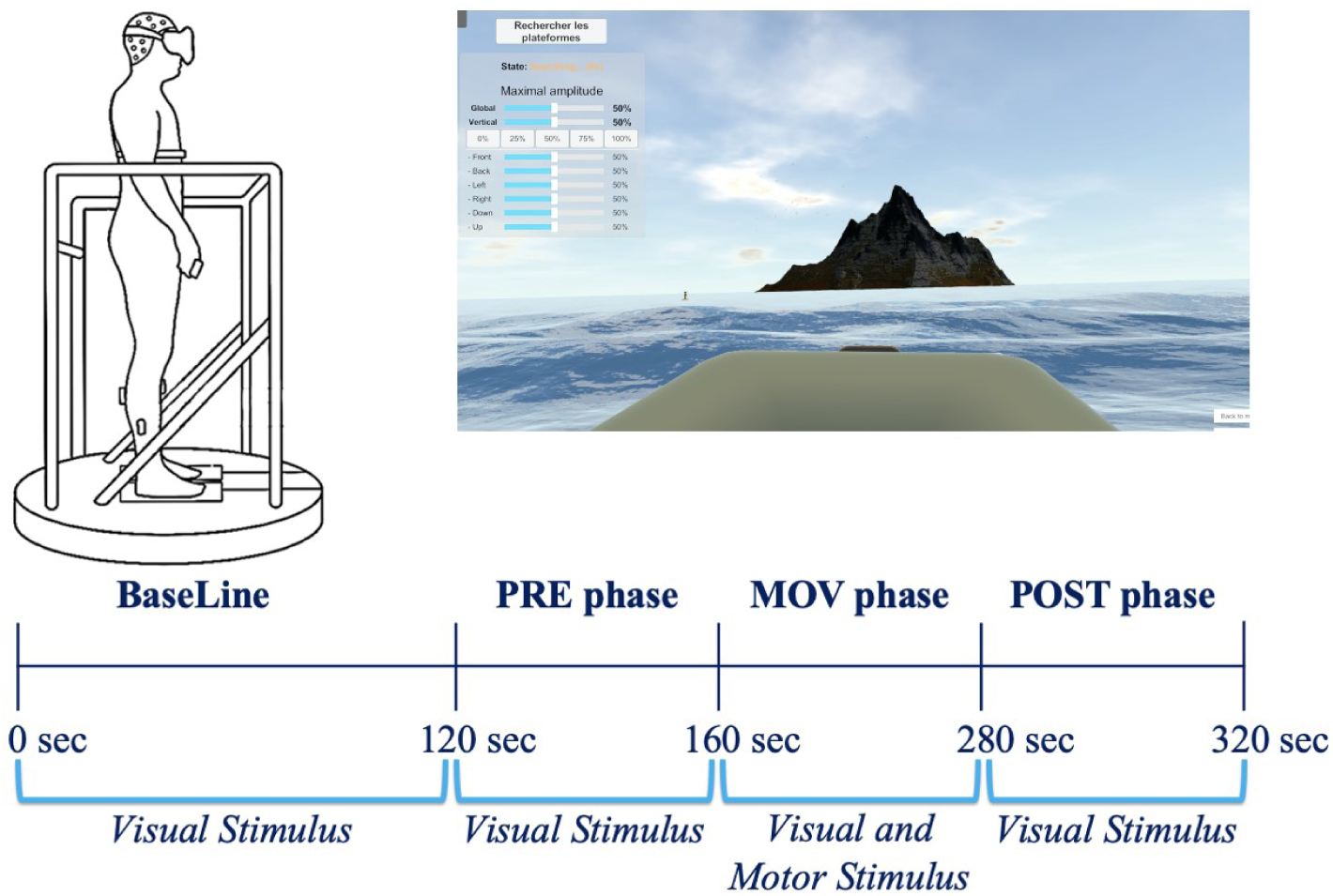
BioVRSea. The different phases of the simulation are characterized by distinct types of stimulation: Baseline (0-120 sec) with dark mountain view and no visual or motor stimulation; PRE Phase (120-160 sec) with visual simulation of the boat; MOV Phase (160-280 sec) with visual-motor stimulus and the platform moving at different amplitudes; POST Phase (280-320 sec) with only visual stimulus remains after the platform stops moving.

Data acquisition during the experiment is systematically conducted by integrating various measurement systems, including EEG, electromyography, heart rate variability, electrodermal activity, and a force platform. These data are integrated to compare PC between individuals. In this study, we focused exclusively on the EEG data and did not include the other measurements in our comparisons.

The experimental protocol is divided into four main phases, each designed to stimulate a variety of responses. As shown in Fig. 1, the protocol begins with the baseline phase (BL), during which the participant stands on a platform with their arms at their sides for two minutes after the acquisition starts. During this phase, the VR environment displays a mountainous landscape to activate all sensors and prepare the system for the upcoming experiment. This is followed by the pre-motor stimulus phase (PRE), during which visual stimuli simulating ocean waves are presented through the VR goggles. Then, the platform begins to move, initiating the movement-stimulus phase (MOV), during which participants are asked to grasp a bar. The platform executes synchronized movements of varying amplitudes, integrating motor and visual stimuli. The experiment concludes with the post-motor stimulus phase (POST), during which the movement of the platform is suddenly stopped while the VR sea simulation continues. In this phase, participants are required to maintain their balance without holding the bar again, as only visual stimuli are presented. This phase introduces the additional challenge of residual instability from the earlier platform movements. The previously listed phases (PRE, MOV, and POST) each last 40 seconds, bringing the total experiment duration to approximately five minutes.

BioVRSea aims to measure the latency between the pre-motor phase and the post-motor phase following the induction of movement. This comprehensive approach aims to understand the physiological responses elicited by the BioVRSea simulation, providing valuable insights into complex PC tasks under conditions of sensory conflict.

### Questionnaire

Before the experiment, each participant completes a questionnaire designed to gather information about their physical condition, neural status, lifestyle, and family history. Additionally, participants answer questions inspired by Golding’s Motion Sickness Susceptibility Questionnaire (MSSQ) [50] to assess past MS symptoms. Symptoms such as general discomfort, headache, increased salivation, sweating, and dizziness are used to calculate the BioVRSea Effect Index (BVSEI), which measures changes in symptoms before and after the VR simulation. This index classifies individuals into two groups: those who experience one or more symptom changes after the simulation and those who do not experience any changes [51]. Understanding these symptom changes is particularly relevant for individuals with neurodegenerative conditions, as these diseases can impact postural stability and exacerbate symptoms during tasks requiring balance. The Lifestyle Index is calculated based on responses regarding BMI, physical activity, and consumption of nicotine, caffeine, and alcohol, all of which can influence both overall health and the management of neurodegenerative diseases. Further, two more indices are established: the physiological/Vegetative (PHY) and Neurological/Muscle Strain (NEURO) indices [51]. Both indices are calculated based on the difference in symptoms before and after the experiment. The Physiological/Vegetative Index is based on responses regarding general discomfort, increased salivation, sweating, and nausea, while the NEURO index focuses on general discomfort, fatigue, headache, blurred vision, and dizziness. These indices are crucial for evaluating the effects of the VR simulation on the PC of the participants, especially for those with neurodegenerative diseases that can impair their ability to maintain balance and coordination.

### Data acquisition

EEG data were recorded using a 64-electrode CA-208 wet cap (ANT Neuro, Hengelo, the Netherlands) with a sampling frequency of 4096 Hz, following the 10-20 system for electrode placement (Table 2). Contact between the electrodes and the subjects was made using conductive gel. The gel was applied via a blunt needle syringe to ensure low impedance between the scalp and the electrodes, maintaining levels below 40 kΩ for optimal conductivity. The EEG cap was connected to the amplifier (ANT Neuro, Hengelo, the Netherlands), which interfaced with a tablet for data acquisition.

**Table 2.** EEG cap. Electrode placement across different brain lobes for the CA-208 waveguard cap.

| Lobes | Electrode names |
| --- | --- |
| Frontal | FP1, FPz, FP2, AF7, AF3, AFz, AF4, AF8, F7, F5, F3, F1, Fz, F2, F4, F6, F8, FC3, FC1, FCz, FC2, FC4, C3, C1, Cz, C2, C4 |
| Temporal Left | FT7, FC5, T7, C5, TP7, CP5 |
| Temporal Right | FC6, FT8, C6, T8, CP6, TP8 |
| Parietal | CP3, CP1, CPz, CP2, CP4, P7, P5, P3, P1, PZ, P2, P4, P6, P8 |
| Occipital | PO7, PO3, POz, PO4, PO8, O1, Oz, O2 |

### Data pre-processing

Signal pre-processing was performed using the EEGLAB Matlab toolbox to eliminate artifacts, eye blinking, and line noise. Each signal was manually inspected to customize our pipeline. The EEG signals were down-sampled to a frequency of 1024 Hz from the original sampling rate. A dual-filter approach was applied, incorporating a low-pass filter at 1 Hz and a high-pass filter at 45 Hz to enhance signal clarity and diminish noise interference. Channels with flat signals or a correlation coefficient below 60% from neighboring channels were excluded from further analysis as noisy channels. The initial step in artifact removal utilized the Artifact Subspace Reconstruction (ASR) method [52]. Then, independent component analysis (ICA) was performed using the probabilistic ICA for reliable data (PICARD) method [53], known for its effectiveness in handling dynamic recordings. Non-brain components were rejected during this step. Afterward, the previously removed and flat channels were interpolated, while the mastoid electrodes (M1 and M2), along with PO5 and PO6, which measure neck activity, were excluded from the analysis. Finally, the remaining channels (excluding EOG) were interpolated back to 61, and the data were re-referenced to the average.

### Data processing: Connectivity analysis to dynamic network computation

Several preliminary steps were required before computing the dynamic brain networks, as described in Fig. 2. EEG source connectivity is a valuable method for identifying brain networks linked to specific cognitive functions, as demonstrated by Hassan et al. [48].

**Fig 2.**
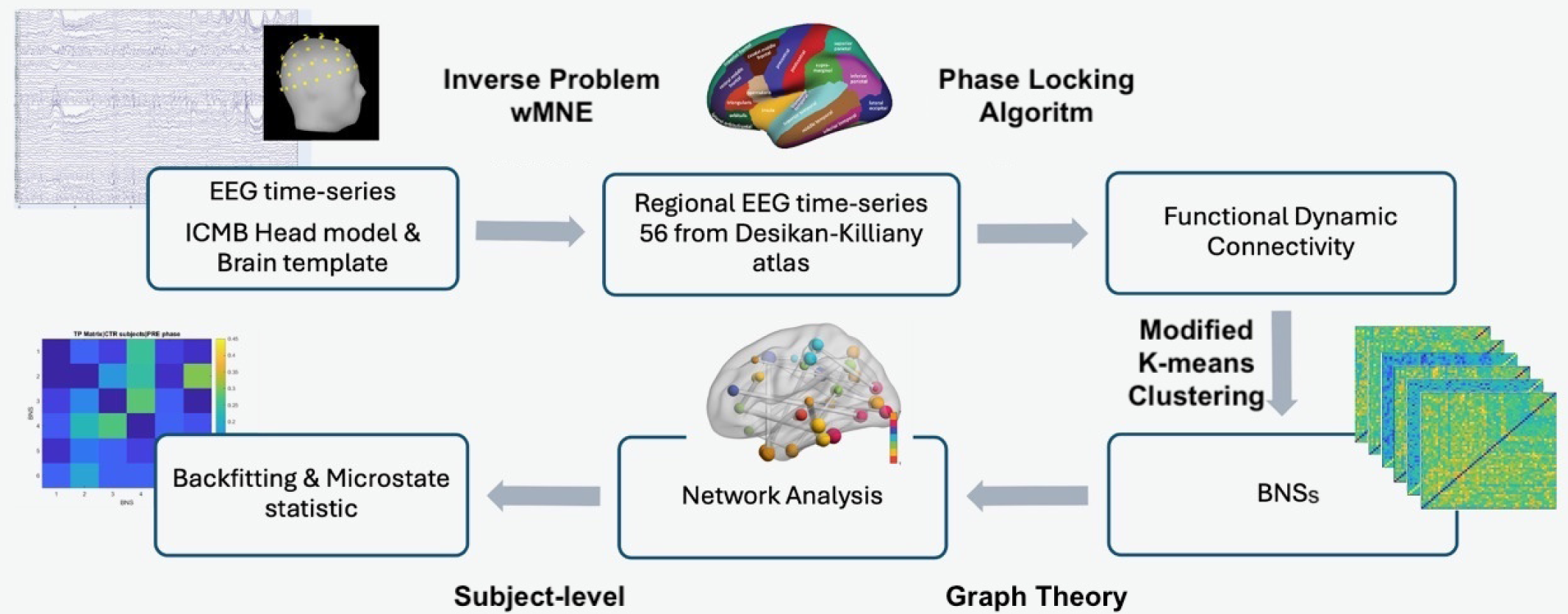
Workflow description. EEGs were recorded using 64 electrodes during the BioVRSea experiment. The cortical sources were reconstructed by solving the inverse problem using the wMNE method. After reconstructing the signals based on brain sources, they were reorganized according to 68 brain ROIs of the Desikan-Killiany atlas. Some areas were aggregated to achieve 56 ROIs, aiming for fewer than the 64 total channels. Functional connectivity matrices were computed between the 56 regional time series using the PLV method across six frequency bands: Delta (0.5–3 Hz), Theta (4–8 Hz), Alpha (8–13.5 Hz), Beta (14–30 Hz), and Gamma (30–90 Hz). Connectivity matrices were first obtained separately for the PD and CTR groups, then concatenated before being input into the k-means++ clustering algorithm. This clustering process identified distinct BNSs, followed by network analysis at the subject level to compute metrics for group comparisons.

The first step was the reconstruction of brain sources from the signals recorded on the scalp. In fact, estimating connectivity directly from the sensor level is limited by low spatial resolution and the effects of volume conduction. These issues can cause false correlations in connectivity measurements, as signals from different electrodes may originate from the same neural source. As a result, connectivity at the sensor level does not accurately reflect functional interactions between brain areas. To address this, we used EEG source localization techniques to reconstruct cortical activity, which involves solving the complex EEG inverse problem. The neural sources for artifact-free EEG data were estimated using the following equation.

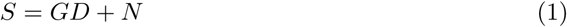

where *S*(*t*) represented the EEG signals recorded at *M* electrodes, *G* is the lead field matrix that models how sources contribute to the scalp potentials, *D*(*t*) denotes the unknown brain activity at *P* source locations, and *N* (*t*) accounts for noise and modeling errors. To solve the inverse problem, we used the *Weighted Minimum Norm Estimate (wMNE)*, given by the formula:

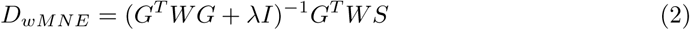

where *G* is the lead field matrix, *W* is a depth-weighting matrix, *λ* is a regularization parameter, and *S* represents the EEG signals.

We used the ICBM MRI template with three-layer segmentation (scalp, outer skull, and inner skull) from the OpenMEEG plugin within the Brainstorm toolbox. The Boundary Element Method (BEM) was employed to model the head volume conductor, validated by prior research [54–56]. The initial 45 seconds of the EEG were used to construct the noise covariance matrix, while the remaining 273 seconds were allocated for source reconstruction. The signals were then divided into the 68 brain regions of interest (ROIs) based on the Desikan-Killiany atlas [57]. Some ROIs were aggregated (aiming at a lower number of ROIs than the number of channels), resulting in a final set of 56 ROIs (Supporting information S1 Table).

Functional connectivity between the EEG time series of the 56 ROIs was calculated across multiple frequency bands (Delta: 0.5-3 Hz, Theta: 4-8 Hz, Alpha: 8-13.5 Hz, Beta: 14-30 Hz, Gamma: 30-90 Hz) using the Phase Locking Value (PLV) algorithm with a sliding window approach (50% overlap and window size of six cycles) [58]. Together with wMNE, this method performed best during cognitive tasks [48].

The PLV computation produced 56x56 square dynamic matrices for each band. The number *N*, representing the total number of computed connectivity matrices, is detailed in Supporting information (S1 Appendix). Specifically, for the alpha band, 953 matrices were created. Each matrix element *A_i,j_* represented the strength of the connections from node *i* to node *j*, referred to as an *edge* in graph theory-based analysis (Table 3).

**Table 3.** Dynamic Connectivity Variables: A list of the number of matrices (*N*) calculated for each frequency band. The dimension (*n*) of each matrix is based on the number of ROIs, with each element (*A_i,j_*) representing the connectivity between the respective regions.

| Variable | Description | Band | Value |
| --- | --- | --- | --- |
| N | Number of dynamic connectivity matrices | Alpha | 953 |
|  |  | Beta | 1953 |
|  |  | Delta | 226 |
|  |  | Gamma | 3407 |
|  |  | Theta | 544 |
| n | Dimension of dynamic connectivity matrices | 56x56 |  |
| $A_{i,j}$ | Element of one connectivity matrix | — | |

BNS segmentation was performed on this dataset using a clustering algorithm to identify stable periods over time. The analysis was based on functions from the EEGLab Microstate toolbox [59], originally designed for EEG voltage topography. We adapted the microstate analysis pipeline described by Poulsen et al. to extract BNSs from functional connectivity data [59]. The optimal number of classes was determined based on goodness-of-fit metrics, including Global Explained Variance (GEV) criterion [59, 60].

For the clustering process, we transformed the symmetric connectivity matrices into vectors of size (*n ×* (*n −* 1)*/*2) = 1540 by extracting the upper triangular elements, excluding the diagonal. Each vector was then concatenated across all time points, resulting in a subject-specific matrix of dimensions 1540 *×* 953, where each column represents a time point, and each row corresponds to a specific connection within the network. These matrices were concatenated among subjects and served as input for the Modified K-means (k-means++) clustering algorithm [61], which has been successfully used in other previous studies on the dynamic functional connectivity of EEG and fMRI [62, 63]. Clustering was performed with the following parameters: the number of clusters varied from 2 to 10, the number of random initializations (rep) was set to 100, and the maximum number of iterations (iter) was 1000, with a default convergence threshold of 10*^−^*^6^. A meta-criterion was applied to obtain the optimal number of clusters [62, 64–66], which was determined to be six. Each identified cluster was defined as a BNS, representing the temporal evolution of brain connectivity. K-means++ was run multiple times with different parameters to ensure the robustness of the clustering results. The BNSs obtained from each clustering iteration were then compared using Pearson correlation analysis to assess the consistency of the identified patterns. A strong correlation indicated high similarity between BNSs across different clustering runs, validating the reliability of the extracted states (Table 4).

**Table 4.**
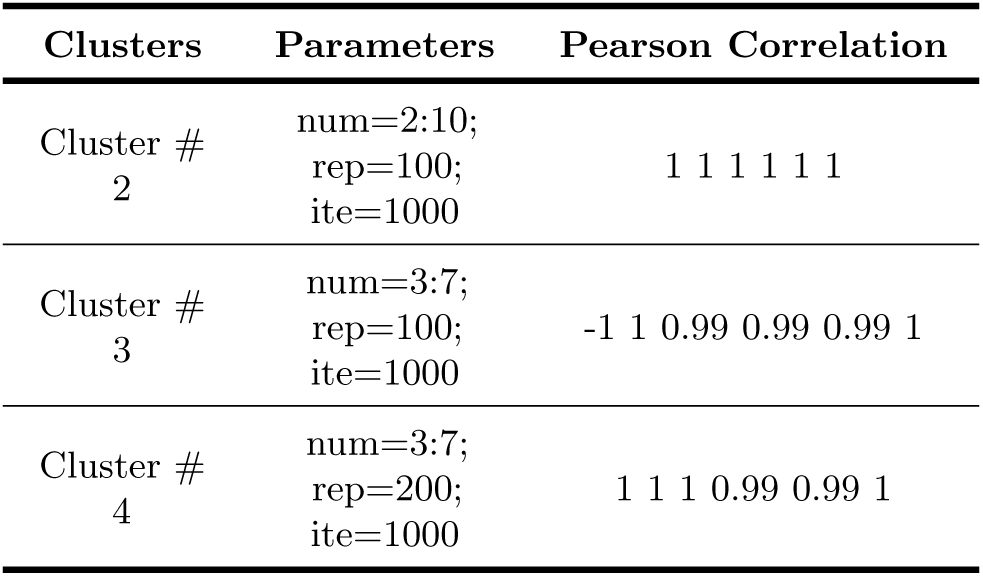
Pearson correlation of BNSs across different clustering runs. Clustering was performed four times with different configurations, varying the number of clusters (num), repetitions (rep), and iterations (ite). Pearson correlation (max = 1) measures BNS equivalence across runs, confirming stability across most conditions.

The topological visualization of the six BNSs was conducted, and they were labeled based on the most prevalent RSNs, as detailed in Subsection “Data processing: Association of BNSs with RSNs”. Each BNS was characterized by distinct spatial distributions and temporal dynamics that correspond to specific cognitive and functional processes. This allowed for a more intuitive interpretation of the BNSs in relation to established RSNs, facilitating comparisons between the two groups in our study.

As a final step, we computed the back-fitting at the subject level using Global Map Dissimilarity (GMD) to identify the best-fitting BNSs throughout the experiment. The smoothing process involved rejection based on small-segment criteria of five matrices [40, 59,60]. For the identified BNSs, we calculated several common metrics used in microstate analysis, including Global Field Power (GFP), occurrence per second (occurrence), average duration (duration), percentage of time occupied (coverage), global explained variance (GEV), and the transition probabilities (TP). These metrics were subjected to statistical analysis between the two groups. A comprehensive description of these metrics is provided in Supporting information (S2 Table).

### Data processing: Association of BNSs with RSNs

Each obtained BNS was computed from a vector to form a 56 × 56 symmetric matrix, that represented the associated functional network. Based on the studies [45, 67], each of the 56 nodes was assigned to one of seven RSNs: Default Mode Network (DMN), Dorsal Attention Network (DAN), Salience Network (SAN), Motor Network (MOT), Auditory Network (AUD), Visual Network (VIS), and Other. The node-RSN affiliations are detailed in Supporting information (S1 Table). Using the Brain Connectivity Toolbox [68], the strength of each node was calculated, representing the sum of the weights of all links connected to that node. Subsequently, we determined the strength of each RSN by summing the values of nodes associated with the specific RSN and normalizing this sum by dividing it by the number of nodes within the RSN. To identify the dominant RSN for each BNS, we considered those with a value exceeding the sum of the average and standard deviation of the strengths of the seven RSNs. We utilized BrainNet Viewer [69] for visualization, where each ROI was represented by a color corresponding to its specific RSN. Each network was displayed with a sparsity of 0.01.

### Correlations between questionnaire scores and brain state metrics (coverage, occurrence, and duration)

The study examined the relationships between coverage, duration, and occurrence variables derived from the BNS analysis of the PRE and POST phases, and the two indices PHY and NEURO. Spearman correlation analysis was conducted to evaluate these relationships. Data were analyzed iteratively for each BNS and subject. Significant correlations (*p <* 0.05) indicated both linear and monotonic relationships across the data subsets.

### Statistical analysis

The statistical analysis was conducted to investigate the effects of the BNSs and subject group on occurrence, duration, and coverage across different phases of the experimental protocol. The study utilized a linear mixed-effects modeling approach (LMM) to account for the hierarchical structure of the data, where repeated measures within subjects were nested within the levels of the BNSs factor. The primary objective was to ascertain any significant main effects and interactions between the subject group (PD vs. CTR) and the BNSs. Model fit was assessed by examining model summary statistics, including coefficients, standard errors, and p-values derived from likelihood ratio tests. Additionally, an analysis of variance (ANOVA) was performed to assess the overall significance of the fixed effects in the model. To further elucidate significant effects identified by the primary analysis, post-hoc pairwise comparisons were conducted using estimated marginal means. These comparisons aimed to delineate differences in the specific metrics between specific levels of the BNSs factor within each subject group across different experiment phases. To mitigate the risk of Type I errors arising from multiple pairwise comparisons, p-values were adjusted using the Benjamini-Hochberg method. This method controls the False Discovery Rate (FDR) and helps maintain the overall integrity of statistical inference. Results were visually represented through empirical cumulative distribution functions and pairs plots to illustrate the distributional properties of the data and highlight significant differences between experimental conditions.

## Results

### BNSs presence

Following the back-fitting process, we initially identified distinct patterns in the presence of BNSs between individuals with PD and CTR subjects across the experimental phases (Fig. 3a, Fig. 3b). All BNSs appeared in each phase for both cohorts, but their relative frequencies varied. Specifically, BNS1 was slightly more frequent in the PD group, except during the POST phase, where it was higher in the CTR group. The trend of BNS1 in the PD group remained consistent across all phases except for the POST phase, whereas in the CTR group, it differed in the PRE phase. BNS2 was consistently less frequent in the PD group across all phases. Conversely, BNS3 and BNS5 were consistently more prominent in the PD group. BNS4 was more frequent in the PD group except during the PRE phase, while BNS6 showed a higher presence in the CTR group except during the PRE phase.

**Fig 3.**
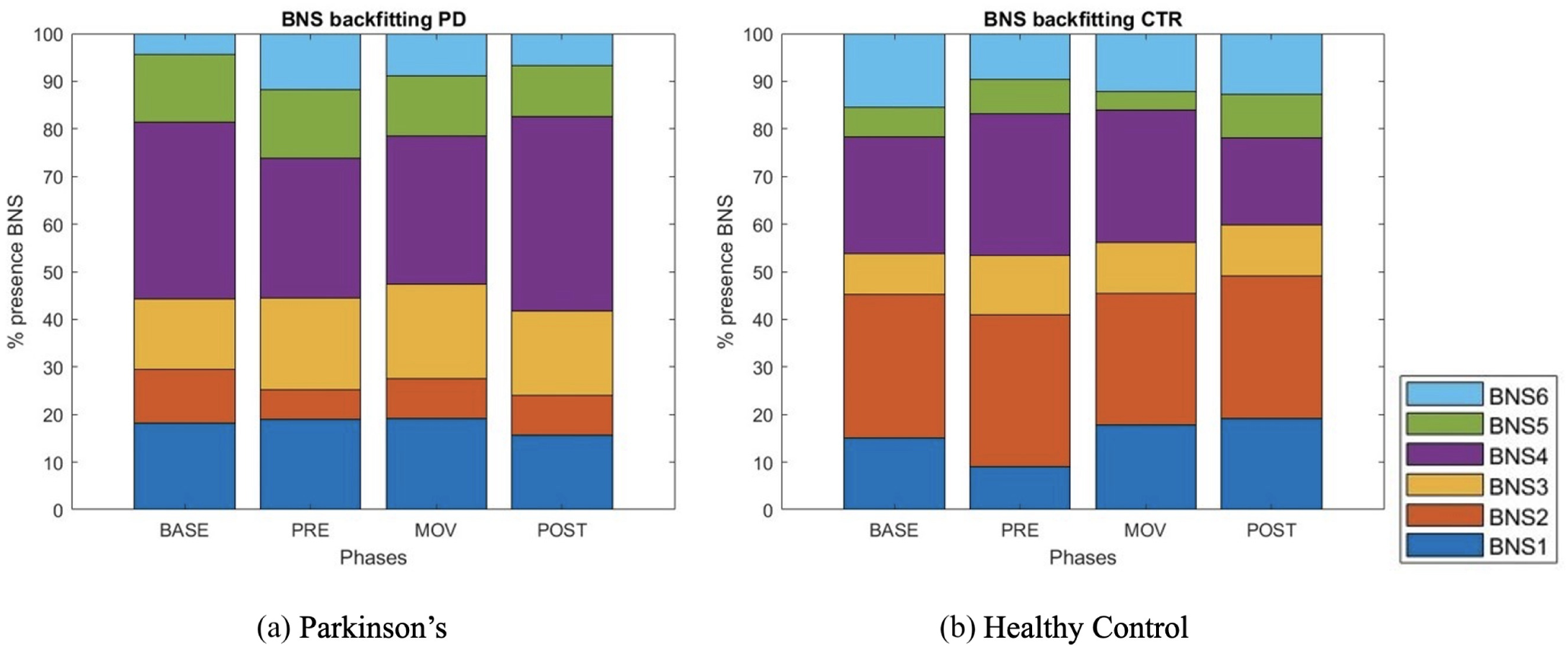
Percentage representation of BNSs. Distribution of BNSs across different experimental time phases for both PD patients and CTRs. Each bar represents the frequency of a specific BNS during the respective phase, illustrating how often each state was engaged throughout the experiment.

### Correlations between questionnaire scores and brain state metrics

The correlation analysis between questionnaire indices and BNS metrics uncovered several significant relationships specific to BNS3. For the CTR group, a notable Spearman correlation was found between the occurrence metric and the NEURO index (*ρ* = 0.27, *p <* 0.01). Both groups showed significant correlations between NEURO scores and the metrics of coverage and duration within BNS3. In the CTR group, the correlation of duration with NEURO was *ρ* = 0.24 (*p <* 0.02), and for coverage, *ρ* = 0.26 (*p <* 0.01). Similarly, in the PD group, the duration-NEURO correlation was *ρ* = 0.21 (*p <* 0.05), and the coverage-NEURO correlation was *ρ* = 0.22 (*p <* 0.05). These findings indicated that both groups demonstrated significant associations between the NEURO index and the metrics of duration and coverage. However, the occurrence metric exhibited a unique correlation pattern that appeared only within the CTR group.

### Statistical analysis of BNSs

We investigated the following parameters of the BNSs: occurrence, duration, coverage, and TP. Statistical analysis revealed significant variations in BNS2 coverage during the PRE (p = 0.019) and MOV (p = 0.036) phases following FDR correction with *α <* 0.05 (Fig. 4), consistent with an initial trend observed in these phases. For BNS4, a similar trend was observed in the BL and POST phases, although it did not remain significant after correction ( S3 Table). No statistically significant results were observed for occurrence; however, a trend was evident in BNS2 during the PRE and MOV phases ( S4 Table). For duration, no statistically significant results were found; however, a trend was noted in BNS5 during the BL phase and in BNS4 during the POST phase. TP analysis indicated that, in individuals with PD, transitions from BNS1 to BNS4 and from BNS4 to BNS1 consistently exhibited significantly higher probabilities compared to the CTR group. Conversely, transitions from BNS4 to BNS1 and BNS3 to BNS4 in CTR subjects displayed higher probabilities than those in the PD group starting from the PRE phase. In PD, activation within the same BNSs occurred later during the POST phase (Fig. 5, S5 Table).

**Fig 4.**
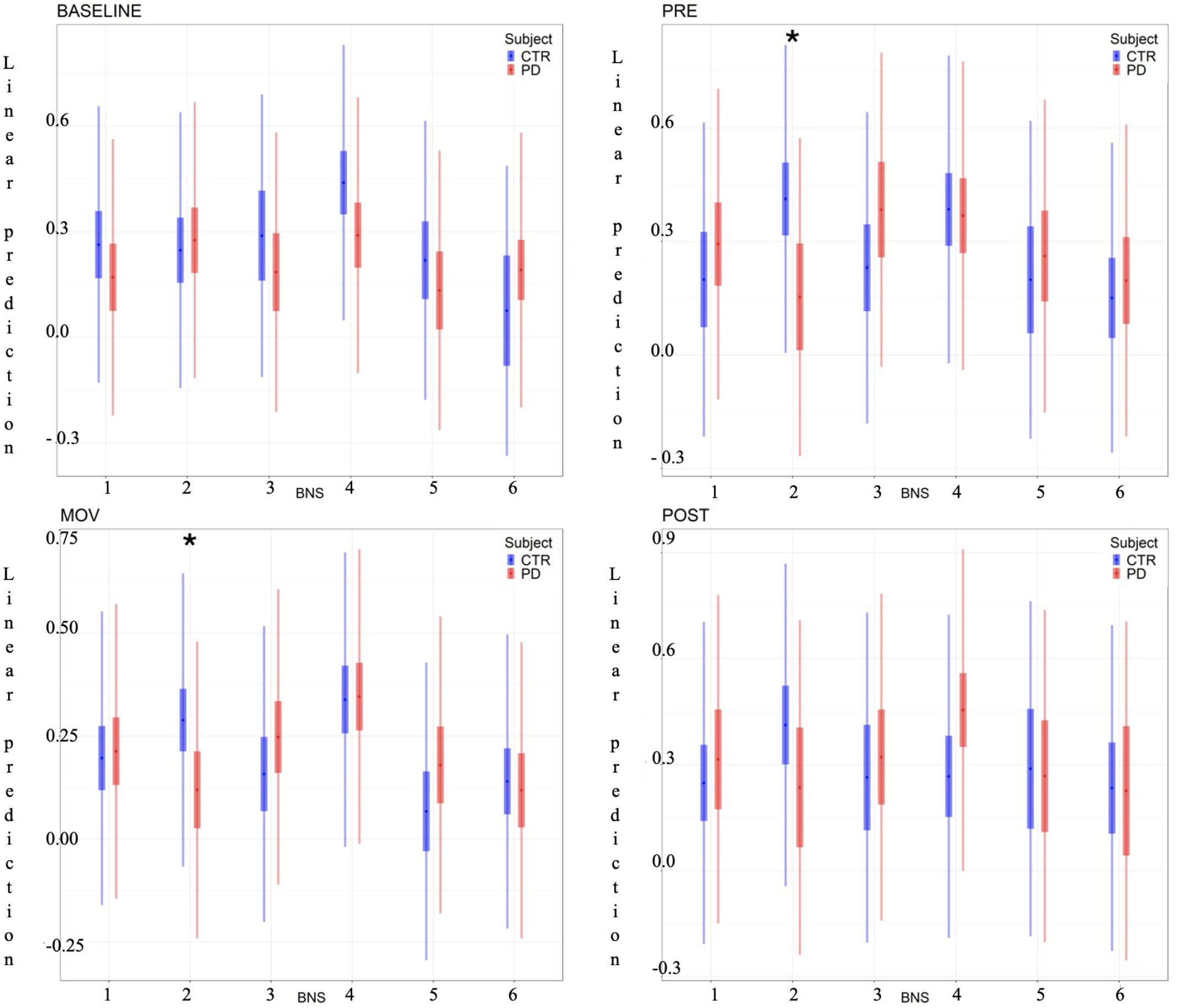
LMM analysis. Coverage for each BNS across all experimental phases in PD patients and CTR. Coverage refers to the proportion of total activity distributed across the six identified BNSs, indicating how dominant each BNS was relative to the others during each phase. Significant differences between groups, corrected using the False Discovery Rate (FDR) method, are highlighted: * p *<* 0.05

**Fig 5.**
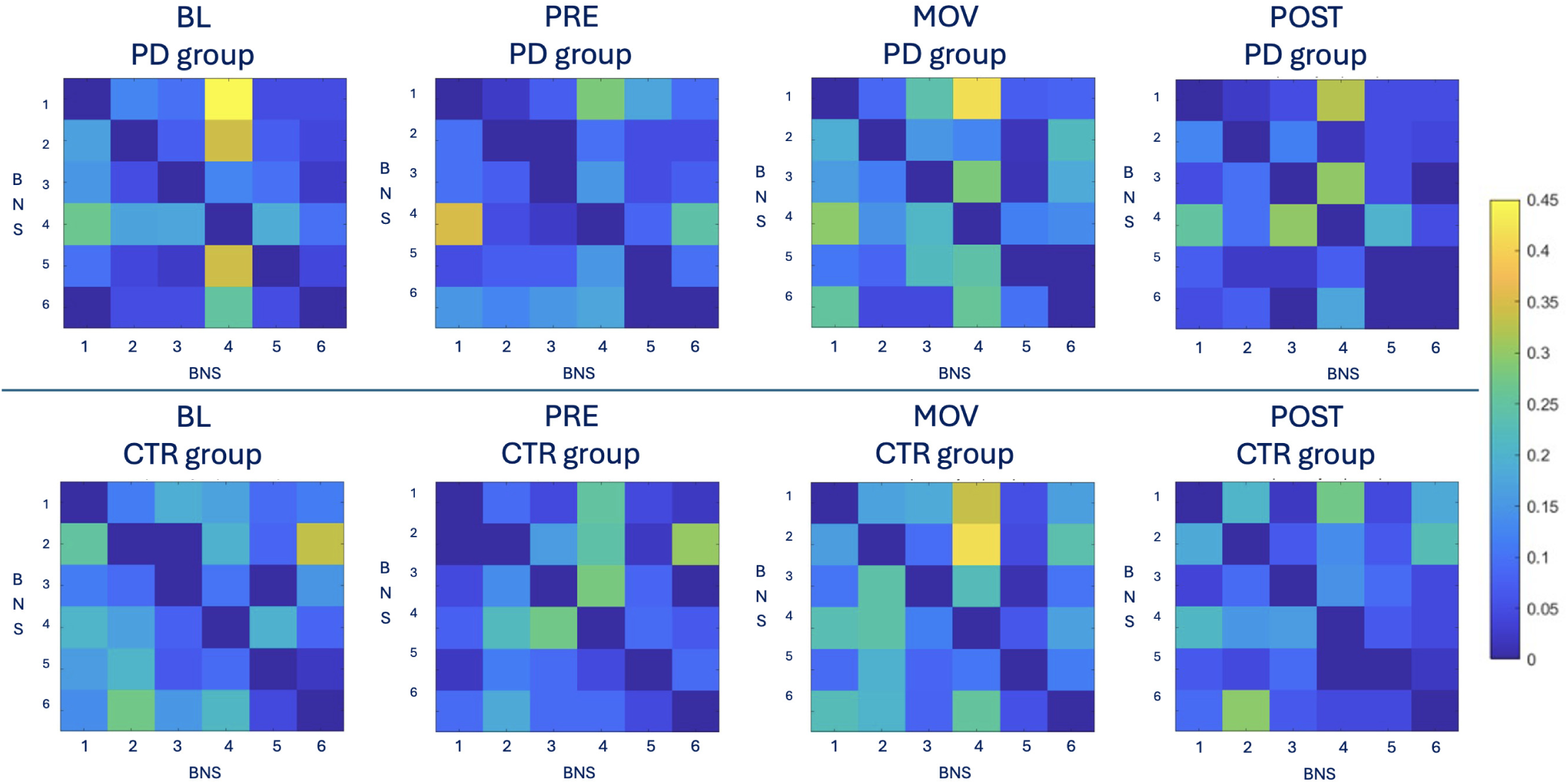
Transition Probability matrices. PD and CTR subjects across BL, PRE, MOV, and POST phases. Each heatmap represents the likelihood of transitioning between different BNSs within each phase, with lighter colors indicating higher transition probabilities. These matrices highlight the patterns of state transitions and the differences between groups in terms of network dynamics.

### Association between BNSs and RSNs

In Fig 6, only the nodes were displayed, where the size of each node represented its strength, corresponding to the overall connectivity at that node. For visualization purposes, a sparsity threshold of 0.01 was applied, displaying only the strongest connections. This approach highlighted the most significant interactions, using color-coding to distinguish different RSNs. In BNS1, the VIS predominated, with nodes concentrated in the left hemisphere, particularly in the parietal and occipital lobes. The SAN was also prominent, with nodes distributed across the frontal and right parietal lobes. In the remaining five BNSs, the DAN was the dominant RSN. BNS2 featured the DAN primarily in the parietal lobe, with nodes of similar sizes spread across the interhemispheric fissure, followed by the SAN in the frontal lobe. BNS3 included the DMN in the frontal lobe, with balanced node strength between the left and right hemispheres, and the AUD in the parietal lobe. BNS4 comprised the DMN, with node strength balanced across the interhemispheric fissure in the parietal lobe, while being exclusively present in the right hemisphere in the frontal lobe. Additionally, the VIS network was mainly localized in the left occipital lobe for the BNS4. BNS5 was similar to BNS4, with larger node sizes across the interhemispheric fissure and in the frontal lobe for the DAN, whereas the SAN was present in both hemispheres across the interhemispheric fissure. Finally, BNS6 involved both the DMN and the SAN, similar to BNS5, but with the DMN absent in the right hemisphere of the parietal lobe.

**Fig 6.**
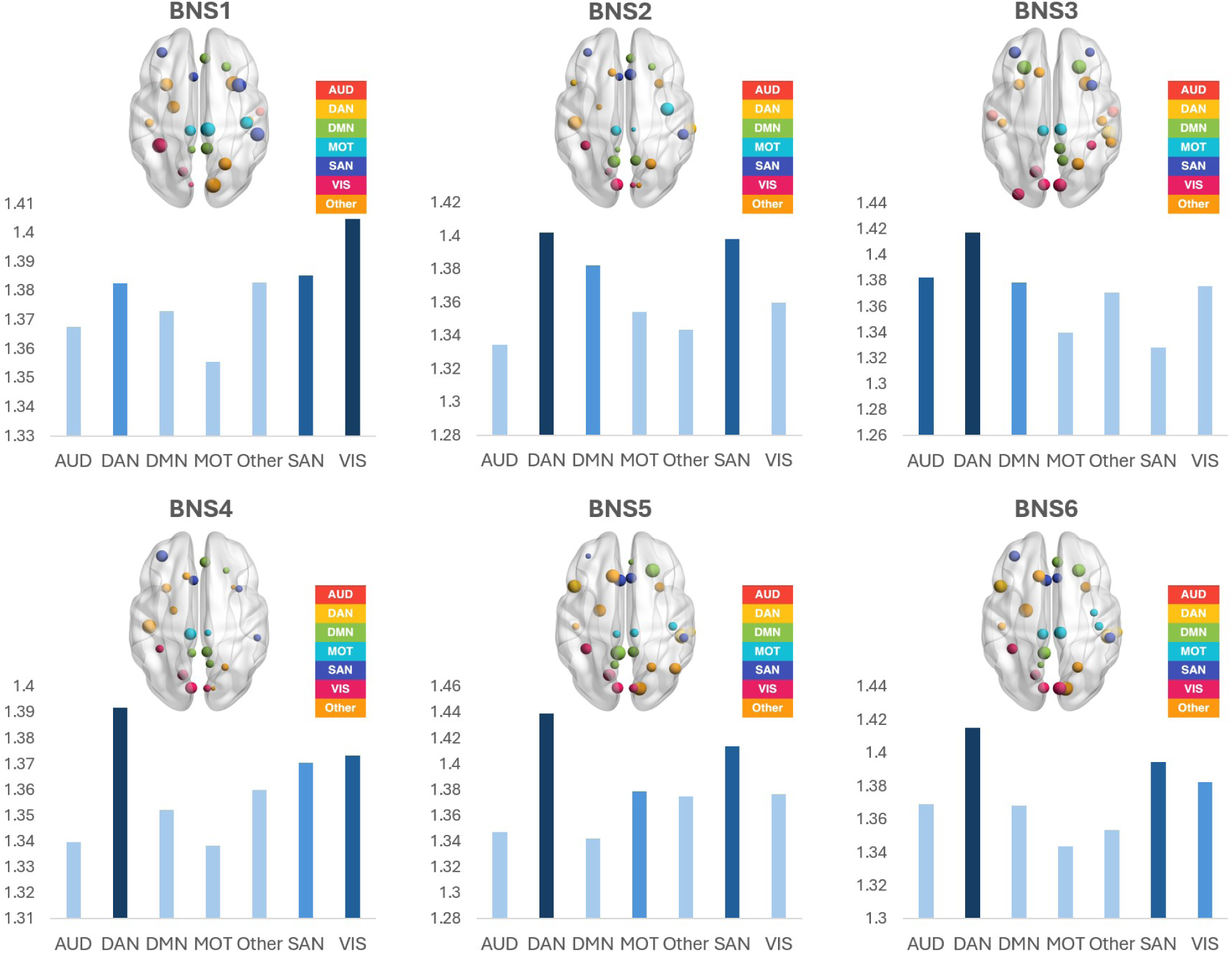
Prevalence of RSNs in each BNS. Calculated as the sum of the node strength values for each ROI associated with the specific RSN. Each BNS is visualized with nodes sized proportionally to their strength (the sum of connections at each node), and edges are shown based on their connectivity, with a sparsity threshold of 0.01 applied to filter weak connections. Different colors represent distinct RSNs, facilitating clear visual differentiation of the dominant networks in each BNS. This visualization highlights the key RSNs contributing to the overall network dynamics across experimental phases.

## Discussion

This study examined brain dynamics during a complex PC task using the BioVRSea paradigm in early-stage PD patients. The novelty of this research lies in its visuomotor stimulation approach, which integrates VR and EEG to investigate motor-cognitive interactions in PD. This allowed for a detailed evaluation of PC impairments and their neural basis. To uncover brain network dynamics, we applied an innovative method adapted from Poulsen et al. [59] to extract BNSs from functional connectivity data. This process included the selection of a clustering algorithm, BNS optimization, and back-fitting to derive key metrics such as coverage, occurrence, and duration.

By analyzing BNSs within the specific BioVRSea paradigm, we aimed to identify new neurophysiological markers of PD that could aid in early diagnosis.

### BNS-Specific analysis

BNSs and RSNs reflected patterns of functional connectivity between different brain regions. While RSNs were typically observed during rest, BNSs represented brain network configurations that occurred during various cognitive tasks or conditions. BNSs involved brain regions that were part of well-known RSNs, such as the default mode network or the fronto-parietal network. The brain states identified through EEG corresponded to RSNs observed in the resting-state. This suggested that certain brain regions remained functionally connected both during rest and while engaging in active tasks. The difference between the presence of RSNs and BNSs in PD compared to CTR referred to alterations in brain connectivity patterns observed in these two groups. This demonstrated altered connectivity and reduced dynamic flexibility of brain networks, refleting difficulties in adapting to cognitive tasks and changing conditions, and highlighted the cognitive and motor impairments associated with the disease.

#### BNS1

BNS1 showed a similar trend in PD compared to CTR, with the only difference in the PRE phase, where the visual stimulation began. Within BioVRSea, BNS1 had the VIS and SAN as its prevalent RSNs (Fig. 3, Fig. 6). The VIS comprised the primary visual cortex (V1) in the striate cortex (Brodmann area 17) and multiple extrastriate regions in the occipital lobe. Together, they covered a significant portion of the posterior cortical surface. This network was essential for processing visual information and played roles in object recognition, motion perception, and spatial awareness tasks. Understanding VIS helped to comprehend visual perception and cognition and to develop treatments for visual disorders and neurological conditions that affect vision [70–72]. A study by Yao et al. [73–75] identified significant differences in occipital function between PD patients without visual hallucinations (PDnonVH) and CTR subjects. The PDnonVH group exhibited a lower amplitude of low frequency fluctuations (ALFF) in the occipito-parietal region, indicating a reduced intrinsic activity in primary and associative visual areas. Additionally, PDnonVH patients showed decreased occipital functional connectivity with other brain regions, suggesting impaired visual processing and integration. The role of VIS and SAN in BNS1 could be more explicitly linked to maintaining alertness and responding to visual and motion cues within BioVRSea. During the PRE phase, when visual stimulation began, SAN activation likely represented participants’ initial engagement with the immersive VR environment, preparing them for the motor challenge introduced in the MOV phase. Highlighting how VIS and SAN networks aligned with BioVRSea task requirements provided important context to their prevalence in BNS1, reflecting the overall cognitive load imposed by the simulated environment in each phase.

#### BNS2

BNS2 was more prevalent in the CTR cohort (Fig. 3), with a notable presence of the DAN and SAN as its primary RSNs (Fig.6). Recent research showed decreased DAN and SAN in PD patients [76, 77]. As shown in Fig. 6, the DAN was prevalent in the rest of the five BNSs. This network was prominent in the brain, primarily consisting of regions that activated during goal-directed tasks. When a specific task or goal was the focus, the DAN directed attention accordingly. It was crucial in tasks requiring sustained attention and cognitive control [70, 78]. Specifically, the DAN was involved in the voluntary deployment of attention, guiding the allocation of attention to specific locations or objects in a top-down manner. In the BioVRSea paradigm, which involved dynamic visual and motor challenges, DAN activation helped participants concentrate on specific postural adjustments to maintain balance, especially under VR conditions where immersive, shifting visuals demanded constant recalibration. In PD, studies indicated that there was a lower percentage of significant connections and reduced connection strength within the DAN compared to controls [76, 79, 80]. This diminished presence of BNS2 in PD participants may, therefore, indicated attentional deficits that hindered PC, suggesting that they might have struggled to maintain the sustained focus required by BioVRSea. The SAN was mainly located in the anterior cingulate cortex and played an essential role in vigilance and in responding to salient stimuli. In the context of BioVRSea, SAN likely supported participants’ alertness to continuous visual stimuli essential for timely postural responses. The lower SAN activation observed in PD may have indicated a compromised arousal system, limiting the ability of PD patients to stay fully alert in the demanding VR environment [70, 81, 82]. Overall, the involvement of both the DAN and SAN in BNS2 highlighted how BioVRSea tasks required attentional focus and responsiveness, underscoring the mental effort these tasks demanded, particularly for PD participants.

#### BNS3

BNS3 was strongly associated with the DAN and the AUD (Fig. 6). These networks played an essential role in integrating sensory information, which explained the significance of BNS3 in the PD group (Fig. 3). The AUD was involved in processing sound stimuli, including inputs from the primary auditory cortex (A1) and peripheral regions like the insula and superior temporal gyrus [70, 83]. In BioVRSea, both visual and auditory inputs challenged balance, making the integration of these sensory cues vital for task performance. In the PD group, the activation of BNS3 reflected a need for intensified sensory integration to compensate for potential deficits in PC. The VR environment incorporated simultaneous visual and auditory stimuli, requiring participants to synchronize these inputs to maintain balance effectively. The presence of both the DAN and AUD in BNS3 emphasized the demand imposed by BioVRSea for attention to and processing of multiple sensory inputs. This dynamic integration was particularly significant in PD, as patients may have relied more heavily on compensatory strategies, utilizing auditory and visual feedback to counterbalance sensory integration deficits in dynamic tasks [70, 84, 85].

#### BNS4

A higher percentage of BNS4 was observed in the PD cohort, with a prevalence of the DAN and VIS as RSNs (Fig. 3, 6). Like BNS1, it engaged the VIS, SAN, and DAN to initiate and maintain attention and awareness in response to early visual stimuli. BNS4 higher activation in the PD group may indicated a sustained compensatory reliance on visual processing for spatial orientation as the task progressed. In BioVRSea environment, this reliance on DAN and VIS in BNS4 could reflected PD participants’ efforts to maintain postural stability through continuous visual feedback, as they may found it more challenging to integrate other sensory signals effectively.

#### BNS5 and BNS6

BNS5 showed a higher percentage in the PD cohort, characterized by the DAN and SAN as RSNs, whereas BNS6 was more prevalent in the CTR cohort, featuring the DAN and SAN as RSNs (Fig. 3, 6). In BioVRSea environment, these networks were crucial for maintaining attention, alertness, and responding to task-relevant stimuli—key elements for balancing in a dynamic visual setting. The study by Putcha et al. [86] suggested that in healthy individuals, the SAN and DMN were anti-correlated, meaning that SAN activation reduced DMN activity and vice versa. This balance was disrupted in PD, where SAN dysfunction, likely due to striatal impairments, affected the ability to suppress DMN effectively [86, 87]. In the context of BioVRSea, where cognitive control and postural adjustments were continually required, this disrupted SAN-DMN coupling in PD may have led to less efficient cognitive responses and adaptive control. As a result, the typical anti-correlation seen in healthy individuals may have become a positive or less negative correlation in PD patients. This altered coupling between SAN and DMN in PD was linked to cognitive deficits commonly observed in the disease [86, 88]. The increased presence of BNS5 in the PD group likely reflected a compensatory mechanism, wherein PD participants have relied more on SAN and DAN activation to maintain attentiveness and actively process stimuli during the VR task. In contrast, the prevalence of BNS6 in the CTR group, also involving SAN and DAN but without the same dysfunctional SAN-DMN interaction, might have supported more fluid cognitive control and attention shifts, aligning with the natural neural dynamics of healthy participants during the task. The primary role of DMN involved inward-focused cognitive processes at rest rather than engagement in task-related activities, explaining why it was not identified as primary during BioVRSea task [70, 89, 90]. Since the experiment required sustained attention and responsiveness to external stimuli, the DMN did not play a central role. However, PD patients’ disrupted SAN-DMN interaction could have affected how effectively they suppressed inward-focused thoughts, impacting their cognitive control and task performance in BioVRSea challenging VR setup.

In summary, the shifts and interactions of the different brain network dynamics highlighted the impact of the disease on cognitive function. The coverage parameter indicated that BNS2 was significantly lower in PD compared to CTR in the two phases of visual and visuomotor tasks (PRE and MOV, respectively), as shown in Fig. 4.

### Neurophysiological differences in BNSs

The correlation analysis provided valuable insights into the neurophysiological effects of PD related to the specific BNS3. This network was associated with the AUD and the DAN. The analysis showed that occurrence is significantly linked to the neurological index (NEURO) from the BioVRSea questionnaire only in the CTR group. However, both groups demonstrated meaningful relationships between duration and coverage with NEURO. This suggests that individuals with PD may exhibit unusual brain activity patterns as they try to manage cognitive challenges. Increased activity in these brain networks might reflect how PD patients perceive their neurophysiological state, potentially indicating higher levels of stress or fatigue, which can negatively impact their quality of life. Overall, the relationships between BNS3 and the activity in AUD and DAN highlighted the neurophysiological changes observed in PD patients, as well as the brain compensatory responses in attention and cognition. The information extracted from the questionnaire can provide valuable insights, particularly regarding neurodegenerative diseases that affect balance and PC. Understanding these symptoms and their correlation with brain networks may assist in developing targeted interventions aimed at improving the overall well-being of patients.

### Implications of BNSs in PC task and in PD

These findings highlighted altered BNS dynamics in PD patients, particularly decreased BNS2 coverage (Fig. 4) and modified transition patterns between BNSs (Fig. 5). These alterations reflected deficits in attention, sensory integration, and network flexibility, all of which were essential for successful balance during dynamic tasks. The reduced BNS2 activity in PD patients during visuomotor phases suggested impaired attention and motor integration, while the increased reliance on visual processing networks (BNS4) underscored compensatory strategies in response to these deficits. The transition probability analysis further revealed PD-specific challenges in switching between states, particularly in moving between attentional (BNS1 and BNS4) and sensory-integration states, which may have contributed to PC deficits (Fig. 5).

### Clinical Relevance

The BioVRSea paradigm researched quantitative metrics to evaluate the early onset of PD. The ultimate goal was to identify the most significant features that could be implemented using simple paradigms suitable for clinical settings.

In neurological diseases like PD, the EEG framework used in this study helped identify key brain dysfunctions during balance tasks, especially in sensorimotor and cognitive networks. The EEG analysis could also detect neural changes linked to PD that might not yet have shown up in motor behavior. By examining BNSs from EEG data, clinicians were able to track changes in brain connectivity that reflected problems with PC, providing early signs of PD and allowing for diagnosis and intervention. The BioVRSea paradigm, combined with brain network analysis, helped develop more tailored treatment plans for PD patients.

## Conclusion

This study aimed to use brain dynamics as a tool to identify new biomarkers for detecting neural dysfunction in early-stage PD, especially during our complex PC task. The proposed approach was a robust methodology that utilized functional connectivity to identify specific BNSs that alternated during the required tasks. By analyzing the differences with healthy individuals, this study intended to uncover neural changes associated with PD that might not have been visible using traditional methods.

The study confirmed the effectiveness of the BioVRSea paradigm in exploring the mechanisms of PC. The visual-motor stimuli required the brain to adjust its physiological responses to regulate body position and the rapid changes in brain activity evidenced this adjustment.

The advanced EEG analysis and clustering algorithms identified six distinct BNSs, with the DAN most active in five states, highlighting its role in behavioral performance and adaptation during attention-demanding tasks. The BNSs distribution on the subject level showed some differences in the BioVRSea phases. Notably, BNS2 was significantly lower in PD patients during the visual and visuomotor phases, suggesting specific deficits in the neural mechanisms underlying PC at this particular stage of the disease, particularly in the brain regions associated with attention and sensory-motor integration.

The focused analysis of alpha band activity revealed significant correlations with PC strategies associated with balance maintenance and postural adjustments. PD patients exhibited altered alpha band dynamics, highlighting potential biomarkers for early diagnosis and intervention. Furthermore, by prioritizing connectivity metrics over conventional channel-based approaches, this study provided a more comprehensive understanding of the neural processes involved in PC. The integration of graph theory allowed a detailed exploration of functional connectivity and its impact on PC.

These findings underscored the potential of BNSs dynamics as a diagnostic tool for identifying neural deficits in PD and other neurological disorders. The BioVRSea paradigm and advanced EEG analysis offered a promising approach for developing targeted interventions to improve postural stability in affected individuals. Overall, this study advanced our understanding of brain connectivity dynamics in PC tasks and highlighted the importance of alpha band activity in maintaining balance. The innovative methodologies and findings paved the way for future research and clinical applications to enhance the quality of life for individuals with neural-motor impairments.

## Data Availability

The data are not available as they pertain to individuals with medical conditions and are therefore subject to privacy and confidentiality restrictions. The scripts are available upon request, as the methodology is still in draft form.

## Supporting information

Abbreviations

**S1 Table. ROI-Node-RSN Mapping**

**S1 Appendix. Phase-Locking Value (PLV) Calculation**

**S2 Table. Computational Methods for BNSs metrics.**

**S3 Table. Statistical Result of Coverage.**

**S4 Table. Statistical Result of Occurrence.**

**S5 Table. Statistical Result of Duration.**

## Author Contributions

| Author | Contribution |
| --- | --- |
| Federica Pescaglia | Conceptualization, Data Curation, Formal Analysis, Investigation, Methodology, Software, Validation, Visualization, Writing – Original Draft Preparation, Writing – Review and Editing |
| Lorena Guerrini | Conceptualization, Data Curation, Formal Analysis, Investigation, Methodology, Software, Validation, Visualization, Writing – Original Draft Preparation, Writing – Review and Editing |
| Carmine Gelormini | Formal Analysis, Investigation, Methodology |
| Romain Aubonnet | Conceptualization, Investigation, Methodology, Resources, Supervision, Writing – Review and Editing |
| Gylfi Örn Thormar | Resources |
| Giorgio Di Lorenzo | Conceptualization, Data Curation, Formal Analysis, Supervision |
| Halldór Jónsson jr | Resources, Writing – Review and Editing |
| Mahmoud Hassan | Conceptualization, Methodology |
| Paolo Gargiulo | Conceptualization, Formal Analysis, Funding Acquisition, Investigation, Methodology, Project Administration, Resources, Supervision, Validation, Writing – Review and Editing |

## Acknowledgments

The authors would like to thank the RANNIS, Icelandic Research Fund (Grant of Excellence no: 239612-051), and research fund Landspitali University Hospital.

## Ethics statement

All participants received detailed written information about the study and provided their signed informed consent. The research was conducted in accordance with the principles embodied in the Declaration of Helsinki and in accordance with Icelandic statutory requirements. The study protocol was approved by the Icelandic National Bioethics Committee (no: 17–183-S1 and VSN-20–101).

